# CT-derived Body Composition Associated with Pulmonary Nodule Malignancy and Growth

**DOI:** 10.1101/2024.10.14.24315476

**Authors:** Tong Yu, Xiaoyan Zhao, Grant Kokenberger, Joseph K Leader, Jing Wang, David Xiao, Xin Meng, Michael N Kammer, Eric L Grogan, James Herman, David Wilson, Jiantao Pu

## Abstract

**Objective:** This study investigated the association between body composition and pulmonary nodule malignancy and growth.

**Methods:** A dataset of subjects with indeterminate pulmonary nodules (IPNs) was created from an internal (n=216) and external (n=162) cohort. Five different body tissues were automatically segmented and quantified from baseline and follow-up chest low-dose computed tomography (LDCT) scans using artificial intelligence (AI) algorithms. Logistic Regression (LR) analyses, t-tests, and Person correlation analyses were performed to study the association between body tissues and nodule malignancy, as well as nodule changes such as density, size, and shape. Gender differences were investigated. The area under the receiver operating characteristic curve (ROC-AUC) was used to assess classifier performance. Average feature importance was evaluated using several machine learning models. Causal relationships were analyzed and visualized using a novel directed graph method.

**Results:** Univariate analysis revealed a significant association between *Skeletal muscle density* and nodule malignancy in both genders (p<0.001). The multivariate model based on body composition yielded AUCs of 0.77 (95% CI: 0.71 – 0.84) and 0.63 (95% CI: 0.54 – 0.72) on the internal and external datasets, respectively. The composite model based on body composition and nodule features yielded AUCs of 0.87 (95% CI: 0.82 – 0.91) and 0.62 (95% CI: 0.53 – 0.72) on the internal and external datasets, respectively. S*keletal muscle* and *intermuscular adipose tissue* features were highly ranked among tissue features, with *skeletal muscle density* retaining its highest rank even after adjusting for clinical and nodule features. The causal graph identified two nodule features and *skeletal muscle density* as directly linked to nodule malignancy. *Skeletal muscle density* and *intramuscular adipose tissue density* were identified as nodule growth indicators in both genders.

**Conclusions:** Body composition can serve as a potential biomarker for assessing nodule malignancy and evaluating nodule growth in both genders.

**Summary Statement:** We found that body composition were critical indicators for discriminating malignant nodules from benign ones and for evaluating the nodule growth in both males and females.

**Key Results:**

1. Univariate analysis revealed a significant association between body composition and nodule malignancy in both males and females. Multivariate analysis further demonstrated the predictive ability of body composition features.
2. Feature importance analysis and causal graph analysis identified *skeletal muscle density* as one of the leading features associated with nodule malignancy.
3. *Skeletal muscle density* and *intramuscular adipose tissue density* were identified as nodule growth indicators in both males and females.

## 1. Introduction

Lung cancer is the leading cause of cancer-related mortality worldwide [1], which highlights the need for efficient screening programs to ensure early detection. The American Cancer Society (ACS) recommends annual screening with low-dose computed tomography (LDCT) for at-risk adults [2]. The National Lung Screening Trial (NLST) has shown that LDCT can reduce lung cancer-related mortality by 20% compared to chest radiography (CXR) [3]. A crucial step in the screening process is determining the malignancy of lung nodules detected on LDCT scans. However, LDCT frequently detects lung nodules, 96.4% of which are benign or false positives [4]. This high false positive rate often leads to unnecessary follow-up procedures, including additional imaging and invasive procedures such as biopsies and surgeries, causing anxiety and potential complications for patients.

To differentiate between benign and malignant nodules using CT images, nodule characteristics, such as size, solidness, shape, and density, are widely used by radiologists as the primary indicators of malignancy in clinical practice [5]. A radiologist’s performance heavily depends on their experience with these features, leading to high inter-and intra-reader variability [6]. Consequently, significant effort has been dedicated to developing computational approaches, which generally fall into two categories. The first approach employs traditional machine learning methods, such as support vector machines and random forests [7–9], to identify significant hand-crafted radiographic features similar to those used by radiologists. These features are integrated with patients’ clinical data, such as age and smoking history [7, 10], to classify nodules into different categories. The second approach is based on deep learning (DL) technologies, such as convolutional neural networks (CNN) [11]. DL models require large, diverse datasets with ground truth and lack interpretability because DL models implicitly learn intricate image textures and patterns, functioning as a “black box”.

Body composition has been explored as a potential biomarker for therapy response [12–14] and for predicting survival time and hospitalization [15–17]. However, its relationship with lung nodule malignancy has not been investigated. We believe that body composition could serve as an effective biomarker for assessing lung nodule malignancy and offer insights into the development and progression of lung nodules. The rationale is that body composition can create a conductive environment for tumor growth, which in turn can affect metabolism and, in turn, alter body composition. As the tumor progresses toward malignancy, these changes in body composition may become more pronounced [18].

In this study, we investigated the association between body composition and lung nodule malignancy, evaluating whether the inclusion of body composition features could improve the assessment of nodule malignancy when combined with nodule characteristics and clinical features. We also examined how body composition changes correlate with nodule growth, with particular attention to gender differences. Both statistical and machine learning approaches were employed to identify body composition features associated with nodule malignancy and growth. Additionally, state-of-the-art causal AI technology was utilized to uncover the underlying causal relationships between body composition, lung nodule characteristics, and nodule malignancy.

## 2. Methods and Materials

### 2.1. Study datasets

A dataset of 216 subjects (male: 113 (52.3%), Table 1) from the Pittsburgh Lung Cancer Screening Study (PLuSS) cohort [19] was selected for this study (Supplementary Figure 1). The selection criteria were: (1) the presence of an indeterminate pulmonary nodule on baseline CT scans, and (2) follow-up CT scans available. Cases with small nodules on the baseline CT scans that showed significant growth on the follow-up scans were also included in the study. Nodule malignancy was confirmed either through biopsy or additional follow-up imaging. Follow-up times ranged from 0.30 to 13.19 years, with a median of 2.34 years and a mean of 4.25 years. For each subject, the largest nodule observed on the initial CT scans was involved in analyses. In the 216 pairs of CT scans, nodules size ranged from 4.00 to 25.32 mm in diameter. This study was approved by the University of Pittsburgh Institutional Review Board (IRB 011171).

**Table 1.** Subject demographic and clinical characteristics of the PLuSS training dataset (n=216).

| Variable |  | Malignant (n=120) | Benign (n=96) | Coefficient (95% CI) | P-value |
| --- | --- | --- | --- | --- | --- |
| Age (year) |  | 63.20 (6.38) | 61.51 (7.47) | 0.04 (0.00, 0.08) | 0.076 |
| Gender |  |  |  |  |  |
|  | Male | 61 | 52 | -0.17 (-0.70, 0.37) | 0.543 |
|  | Female | 59 | 44 |  |  |
| Race |  |  |  |  |  |
|  | Non-white | 7 | 5 | 0.12 (-1.06, 1.30) | 0.842 |
|  | White | 113 | 91 |  |  |
| BMI |  | 27.47 (4.07) | 27.67 (4.35) | -0.01 (-0.08, 0.05) | 0.726 |
| BMI category |  |  |  | \ | 0.874 |
|  | < 20.0 | 0 | 2 |  |  |
|  | 20.0 to 24.9 | 34 | 27 |  |  |
|  | 25.0 to 29.9 | 55 | 40 |  |  |
|  | ≥ 30.0 | 31 | 27 |  |  |
| Smoking Status |  |  |  |  |  |
|  | Current | 87 | 59 | 0.50 (-0.07, 1.08) | 0.086 |
|  | Former | 33 | 37 |  |  |
| Cigarette (day) |  | 25.50 (8.49) | 25.21 (8.91) | 0.00 (-0.03, 0.03) | 0.805 |
| Pack (year) |  | 57.88 (21.93) | 53.51 (22.38) | 0.01 (0.00, 0.02) | 0.152 |
| **Emphysema** |  |  |  |  |  |
|  | Yes | 83 | 53 | 0.60 (0.04, 1.16) | **0.036\*** |
|  | No | 37 | 43 |  |  |

Mean (Standard deviation (SD)) and Count for continuous and categorical variables respectively.
Coefficient and p-value from univariate logistic regression model on unstandardized data.
\*, indicates p-value < 0.05.
Values of 0.00 are non-zero but are truncated due to rounding.

An external validation dataset originated from the Vanderbilt University Medical Center (VUMC), denoted as the Vanderbilt dataset [20] (Supplementary Table 1). This dataset consisted of 162 patients (male: 94 (58.0%)) who were enrolled in the VUMC study between 2003 and 2017. These subjects had lung nodules with the largest axial diameter ranging from 6.00 to 30.00 mm. Diagnoses were confirmed through a biopsy or a 2-year longitudinal follow-up imaging showing no signs of growth for benign nodules.

### 2.2. Image acquisition

The chest LDCT scans in PLuSS were acquired at an end-inspiration during a single-breath-hold using a helical technique at 20-40 mA and 120/140 kVp. These images were reconstructed with a high spatial frequency algorithm specifically for lung tissue. The images encompassed the entire lung field in a 512×512 pixel matrix with slice thickness ranging from 1.25 to 2.5 mm. The in-plane pixel dimensions ranged from 0.55 to 0.83 mm. Radiologists reviewed the images using standard lung windows on an imaging workstation, adjusting window width and level as needed to detect nodules and assess mediastinal structures [19].

### 2.3. CT image features

In-house software was used to automatically segment five types of body tissues related to body composition depicted on chest LDCT images (Figure 1(g-j)) [21]. The five tissue types included visceral adipose tissue (VAT), subcutaneous adipose tissue (SAT), intermuscular adipose tissue (IMAT), skeletal muscle (SM) and bones. Volume, mass, and density (i.e., average Hounsfield Unit (HU) value) were computed for each segmented tissue [22].

**Figure 1.**
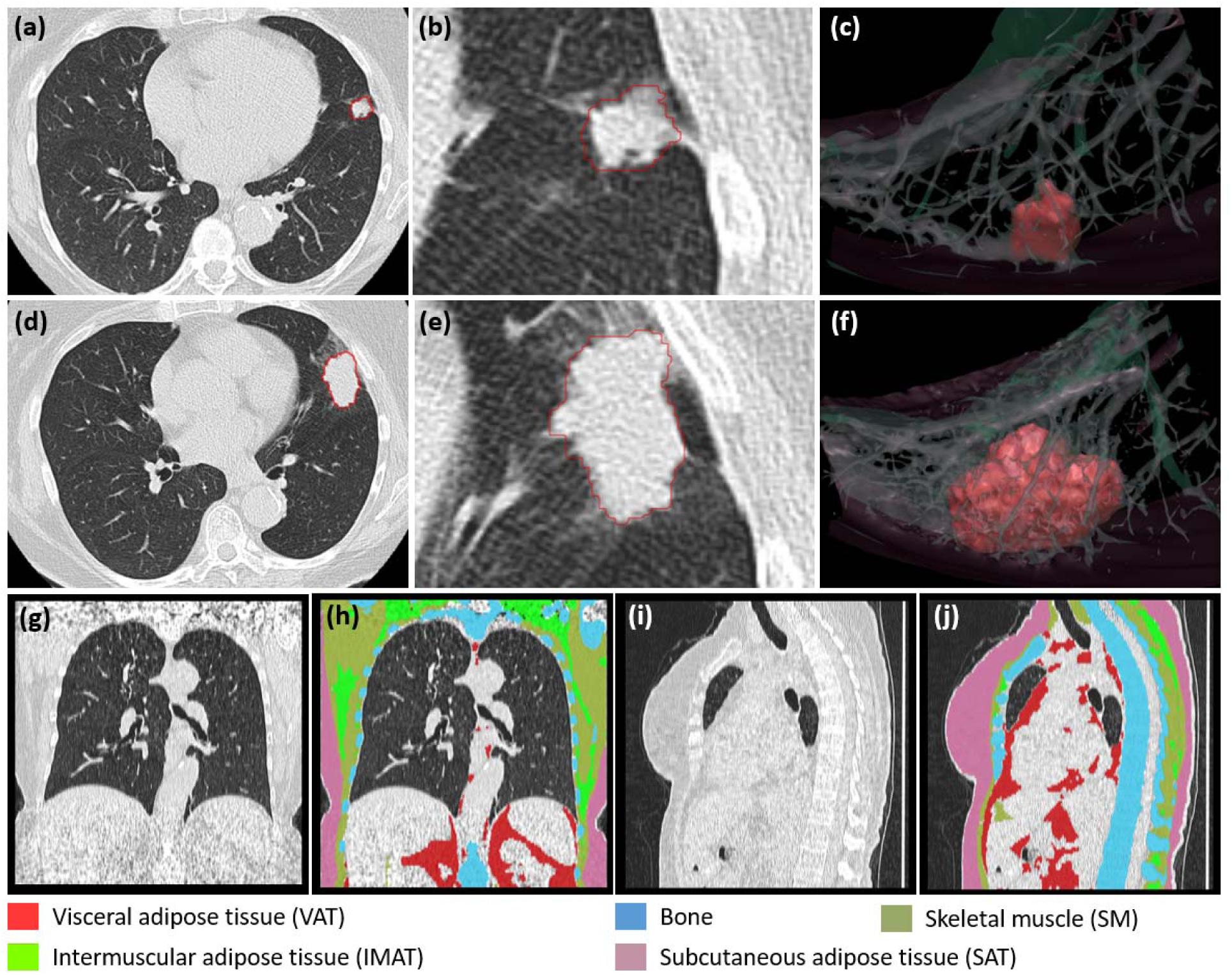
An example illustrating the automated segmentations of lung nodules and five tissues depicted on a LDCT scan with an image thickness of 2.5 mm. (a)-(c): the lung nodule at the baseline CT scan. (d)-(f): the lung nodule at the most recent CT scan. (c), (f): 3-D visualization of the nodules and its surrounding vessels. (g)-(j): tissue segmentation.

Lung nodules were automatically segmented and quantified on chest LDCT images using in-house software [23] based on the following features: 1) volume, 2) mean density, 3) surface area, 4) maximum diameter, 5) mean diameter, 6) mean diameter of the solid portion, 7) solidness, 8) calcification volume, 9) irregularity (Figure 1 (a-f)). Solidness was determined using a threshold of −300 HU. Calcification volume was computed as the volume in the nodule with a HU value greater than 200. Irregularity was computed as the ratio between nodule surface area and volume [24].

### 2.4. Statistical analyses

Logistic regression (LR) was used to analyze the impact of body composition, nodule, and clinical features on lung nodule malignancy using baseline data (from the first CT scan). Univariate LR was implemented on unstandardized data and multivariate LR was implemented on standardized data to classify nodules as benign or malignant. Different multivariate LR models were developed based on body composition, nodule, and clinical features. These models included: (1) *Body composition (BC)* model, (2) *Nodule* model, (3) *Clinical* model, (4) *BC + Clinical* model, (5) *BC + Nodule* model, and (6) *Composite (BC + Clinical + Nodule)*. To construct these models and mitigate potential overfitting, all features were initially included, and then a multi-strategy feature selection approach was applied. First, Least Absolute Shrinkage and Selection Operator (LASSO) regularization was utilized to select features by shrinking and reducing certain LASSO regression coefficients to zero. This procedure was performed using a 10-fold cross-validation (CV) approach to fine-tune the optimal regularization parameter to ensure robustness. Features with LASSO coefficients equating to zero were excluded. Then, variance inflation factor (VIF) selection and backward stepwise selection were applied iteratively. Specifically, features with a VIF larger than a threshold of 3.0 were further removed. This process continued until all remaining features exhibited a p-value less than 0.05.

The performance of the classification models was evaluated in both the model development and external validation datasets using the area under (AUC) the receiver operating characteristic (ROC) curve with 95% confidence intervals (CIs). These CIs were computed using the 10-fold CV method. To compare the performance of two models, the DeLong test was used for independent models, while the likelihood ratio test (LRT) was used for nested models. All statistics were performed in Python 3.11.4 and RStudio 4.3.1.

Body composition indicators associated with nodule growth were analyzed. Changes in body composition and nodules were quantified between baseline and last CT scans. Absolute changes in both body composition and lung nodule features were computed between these two scans. Paired t-tests were used to identify nodule features with significant changes (p < 0.05), and unpaired t-tests were used to compare body composition changes between malignant and benign groups. Pearson correlation analysis was employed to examine the correlation between changes in body composition and nodule features. Person correlation analysis was also used to examine the correlations between baseline body composition features and nodule malignancy univariately.

### 2.5. Feature importance analysis

Feature importance was evaluated using permutation importance (PI) and Shapley Additive Explanations (SHAP) [25]. To model classifiers for nodule malignancy, Support Vector Machine (SVM), Random Forest (RF) and Multi-layer Perceptron (MLP) were used in addition to LR. Feature importance scores provided by each method were standardized using min-max scaling, and then average feature importance scores were calculated. The SVM classifier was developed using a linear kernel with a regularization parameter of 1.0. The RF classifier was constructed with 100 trees and used the Gini impurity criterion. The MLP classifier was trained with 100 hidden layers, a constant learning rate of 0.001, and a maximum of 1,000 iterations until convergence.

### 2.6. Causal relationship analysis

To evaluate the statistical findings and reveal the underlying causal relationships between features, a novel causal discovery method called "Grouped Greedy Equivalence Search" (GGES) [26] was used to generate directed graphs. This allowed the visualization of the causal relationships between clinical features, CT image features, and nodule malignancy. Initially, all features were included in the causal discovery graph, after which only features that have paths leading to the node ’malignancy’ were retained. Features directly connected to the node ’malignancy’ were identified as the most crucial predictive factors for nodule malignancy.

## 3. Results

### 3.1. Body composition at baseline CT scans and nodule malignancy

There were 120 (55.56%) malignant and 96 benign nodules in the PLuSS training dataset. The mean age of the benign and malignant groups were 61.51 (± 7.47) and 63.20 (± 6.38) years, respectively. The results of univariate logistic regression analysis of clinical features, body tissues, and nodule features are presented in Tables 1 and 2. Among the clinical features, only *Emphysema* was found to be significant (p < 0.05). Subjects with emphysema had an increased odds of malignancy of 82.2% (Table 1). Regarding body composition features across all genders, *VAT density, IMAT density, SM density,* and *Bone density*, showed statistically significant negative associations with nodule malignancy (Table 2, Supplementary Figure 2). *SM density* demonstrated the strongest correlation with nodule malignancy (Supplementary Figure 3). When considering gender differences, different body composition features were associated with malignancy (Figure 2; Supplementary Table 2, 3). Specifically, *SM density* maintained a strong negative correlation with malignancy in both males and females, while *bone* features were associated only in males and *VAT* features only in females. Nodule features, including *Mean Intensity, Max Diameter, Mean Diameter, Solidness,* and *Irregularity*, were significantly associated with nodule malignancy in males, females, and all subjects (Table 2, Supplementary Table 4, 5). *Mean Intensity* showed a negative correlation with malignancy, while the other features showed positive correlations.

**Figure 2.**
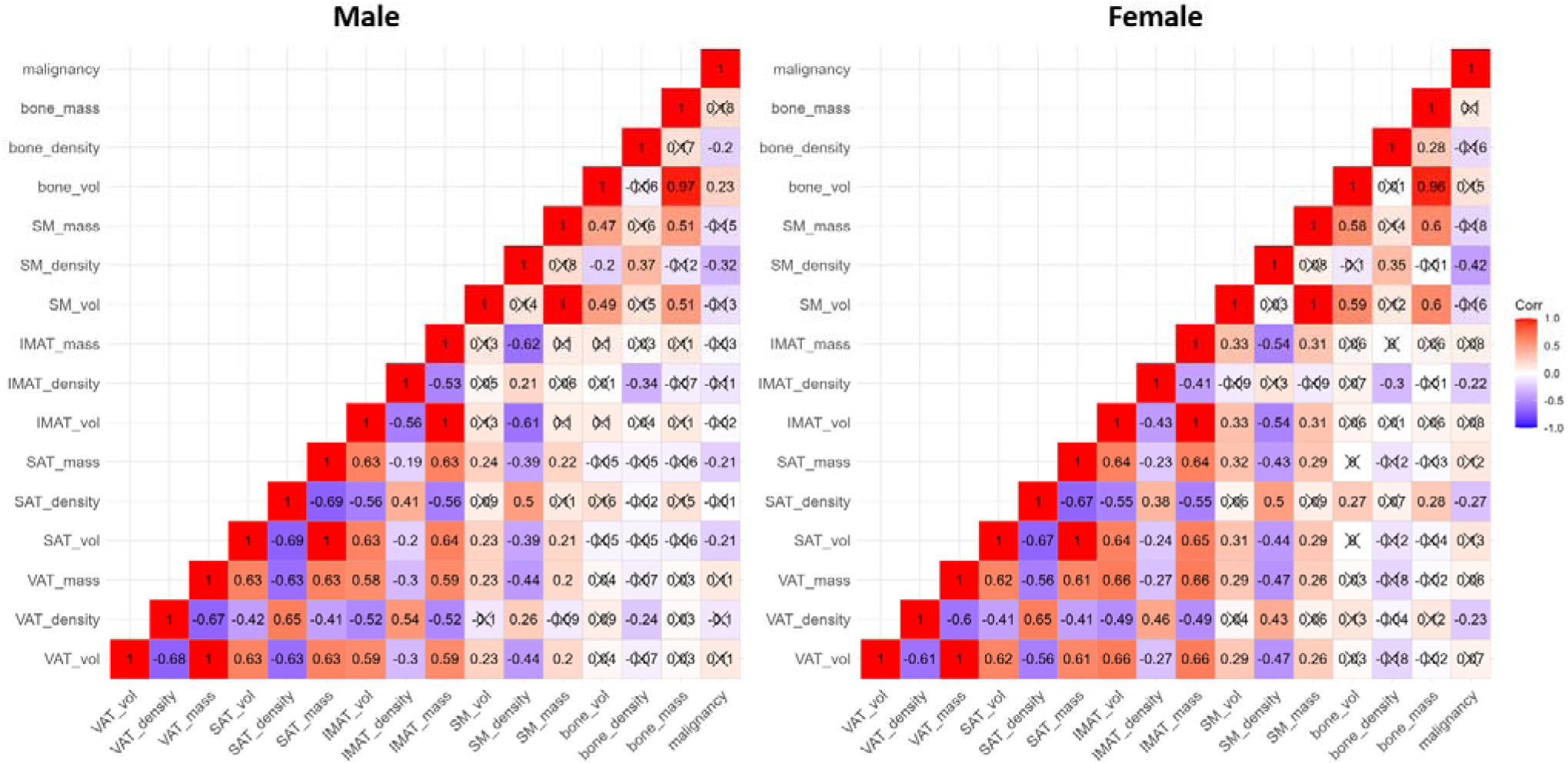
Correlation visualization between baseline body composition and nodule malignancy (Male=113, Female=103). Correlations marked with a cross are statistically insignificant (p-value > 0.05).

**Table 2.**
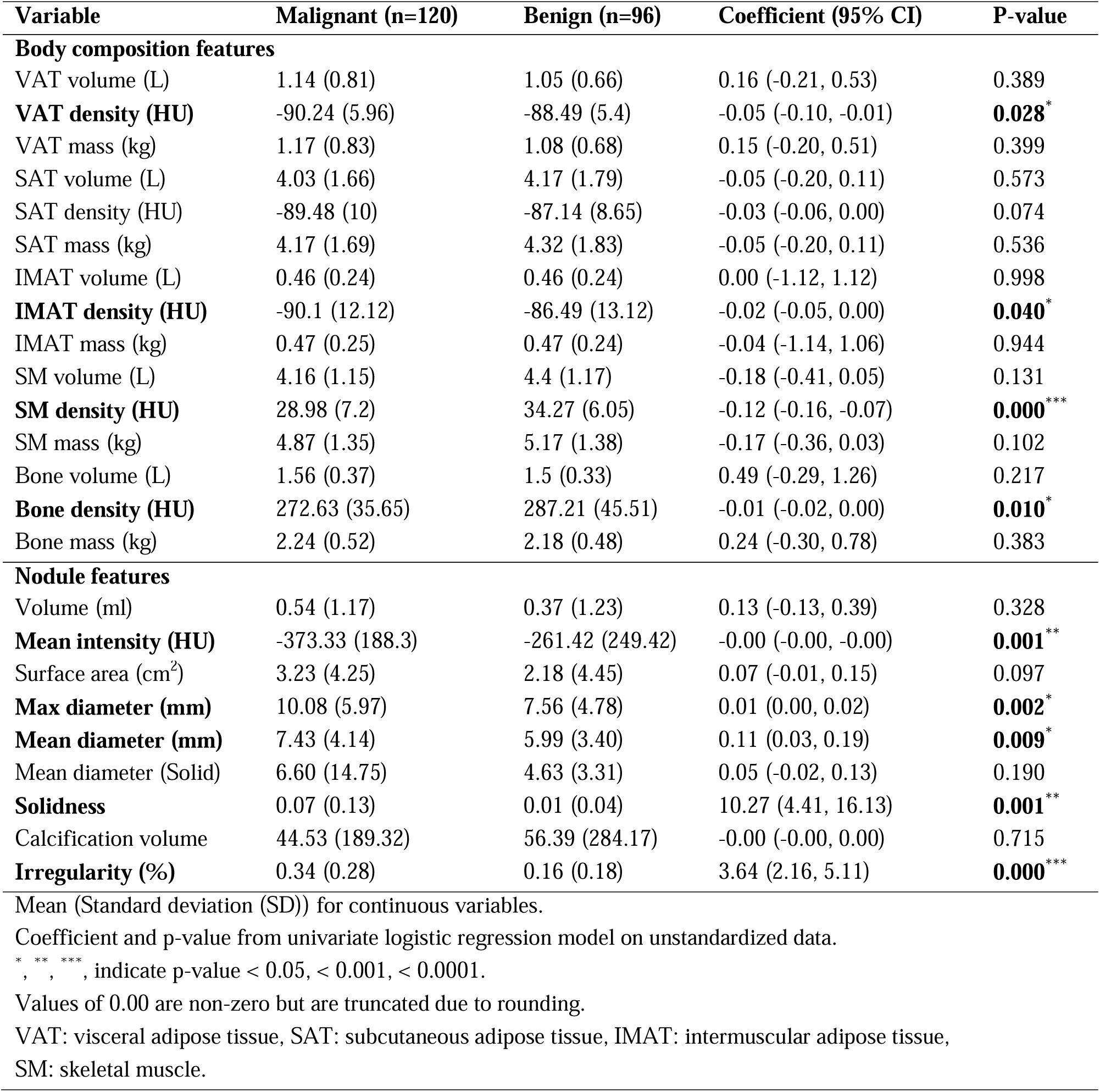
Summary statistics of CT-derived body composition and nodule features in lung cancer and non-cancer groups and their univariate logistic regression results.

| Variable | Malignant (n=120) | Benign (n=96) | Coefficient (95% CI) | P-value |
| --- | --- | --- | --- | --- |
| **Body composition features** |  |  |  |  |
| VAT volume (L) | 1.14 (0.81) | 1.05 (0.66) | 0.16 (-0.21, 0.53) | 0.389 |
| **VAT density (HU)** | -90.24 (5.96) | -88.49 (5.4) | -0.05 (-0.10, -0.01) | **0.028\*** |
| VAT mass (kg) | 1.17 (0.83) | 1.08 (0.68) | 0.15 (-0.20, 0.51) | 0.399 |
| SAT volume (L) | 4.03 (1.66) | 4.17 (1.79) | -0.05 (-0.20, 0.11) | 0.573 |
| SAT density (HU) | -89.48 (10) | -87.14 (8.65) | -0.03 (-0.06, 0.00) | 0.074 |
| SAT mass (kg) | 4.17 (1.69) | 4.32 (1.83) | -0.05 (-0.20, 0.11) | 0.536 |
| IMAT volume (L) | 0.46 (0.24) | 0.46 (0.24) | 0.00 (-1.12, 1.12) | 0.998 |
| **IMAT density (HU)** | -90.1 (12.12) | -86.49 (13.12) | -0.02 (-0.05, 0.00) | **0.040\*** |
| IMAT mass (kg) | 0.47 (0.25) | 0.47 (0.24) | -0.04 (-1.14, 1.06) | 0.944 |
| SM volume (L) | 4.16 (1.15) | 4.4 (1.17) | -0.18 (-0.41, 0.05) | 0.131 |
| **SM density (HU)** | 28.98 (7.2) | 34.27 (6.05) | -0.12 (-0.16, -0.07) | **0.000\*\*\*** |
| SM mass (kg) | 4.87 (1.35) | 5.17 (1.38) | -0.17 (-0.36, 0.03) | 0.102 |
| Bone volume (L) | 1.56 (0.37) | 1.5 (0.33) | 0.49 (-0.29, 1.26) | 0.217 |
| **Bone density (HU)** | 272.63 (35.65) | 287.21 (45.51) | -0.01 (-0.02, 0.00) | **0.010\*** |
| Bone mass (kg) | 2.24 (0.52) | 2.18 (0.48) | 0.24 (-0.30, 0.78) | 0.383 |
| **Nodule features** |  |  |  |  |
| Volume (ml) | 0.54 (1.17) | 0.37 (1.23) | 0.13 (-0.13, 0.39) | 0.328 |
| **Mean intensity (HU)** | -373.33 (188.3) | -261.42 (249.42) | -0.00 (-0.00, -0.00) | **0.001\*\*** |
| Surface area (cm<sup>2</sup>) | 3.23 (4.25) | 2.18 (4.45) | 0.07 (-0.01, 0.15) | 0.097 |
| **Max diameter (mm)** | 10.08 (5.97) | 7.56 (4.78) | 0.01 (0.00, 0.02) | **0.002\*** |
| **Mean diameter (mm)** | 7.43 (4.14) | 5.99 (3.40) | 0.11 (0.03, 0.19) | **0.009\*** |
| Mean diameter (Solid) | 6.60 (14.75) | 4.63 (3.31) | 0.05 (-0.02, 0.13) | 0.190 |
| **Solidness** | 0.07 (0.13) | 0.01 (0.04) | 10.27 (4.41, 16.13) | **0.001\*\*** |
| Calcification volume | 44.53 (189.32) | 56.39 (284.17) | -0.00 (-0.00, 0.00) | 0.715 |
| **Irregularity (%)** | 0.34 (0.28) | 0.16 (0.18) | 3.64 (2.16, 5.11) | **0.000\*\*\*** |

Mean (Standard deviation (SD)) for continuous variables.
Coefficient and p-value from univariate logistic regression model on unstandardized data.
\*, \*\*, \*\*\*, indicate p-value < 0.05, < 0.001, < 0.0001.
Values of 0.00 are non-zero but are truncated due to rounding.
VAT: visceral adipose tissue, SAT: subcutaneous adipose tissue, IMAT: intermuscular adipose tissue, SM: skeletal muscle.

The multivariate logistic regression results of various integrative nodule malignancy models are illustrated in Table 3. The *Composite* model and the *BC + Nodule* model retained the same parameters — body composition features *IMAT, SM*, and *SAT,* and nodule features *Irregularity* and *Mean Intensity* — after feature selection and achieved the best discrimination performance with an AUC of 0.87 (95% CI: 0.82 – 0.91). The *BC + Clinical* model and the *BC* model also maintained the same parameters and exhibited suboptimal performance with an AUC of 0.77 (95% CI: 0.71 – 0.84), incorporating body composition features *IMAT, SM, VAT, SAT*. This was followed by the *Nodule* model, which had an AUC of 0.71 (95% CI: 0.64 – 0.78) (Figure 3a). Clinical features were not included in any of the models, and the *Clinical* model is not presented due to its low performance.

**Figure 3.**
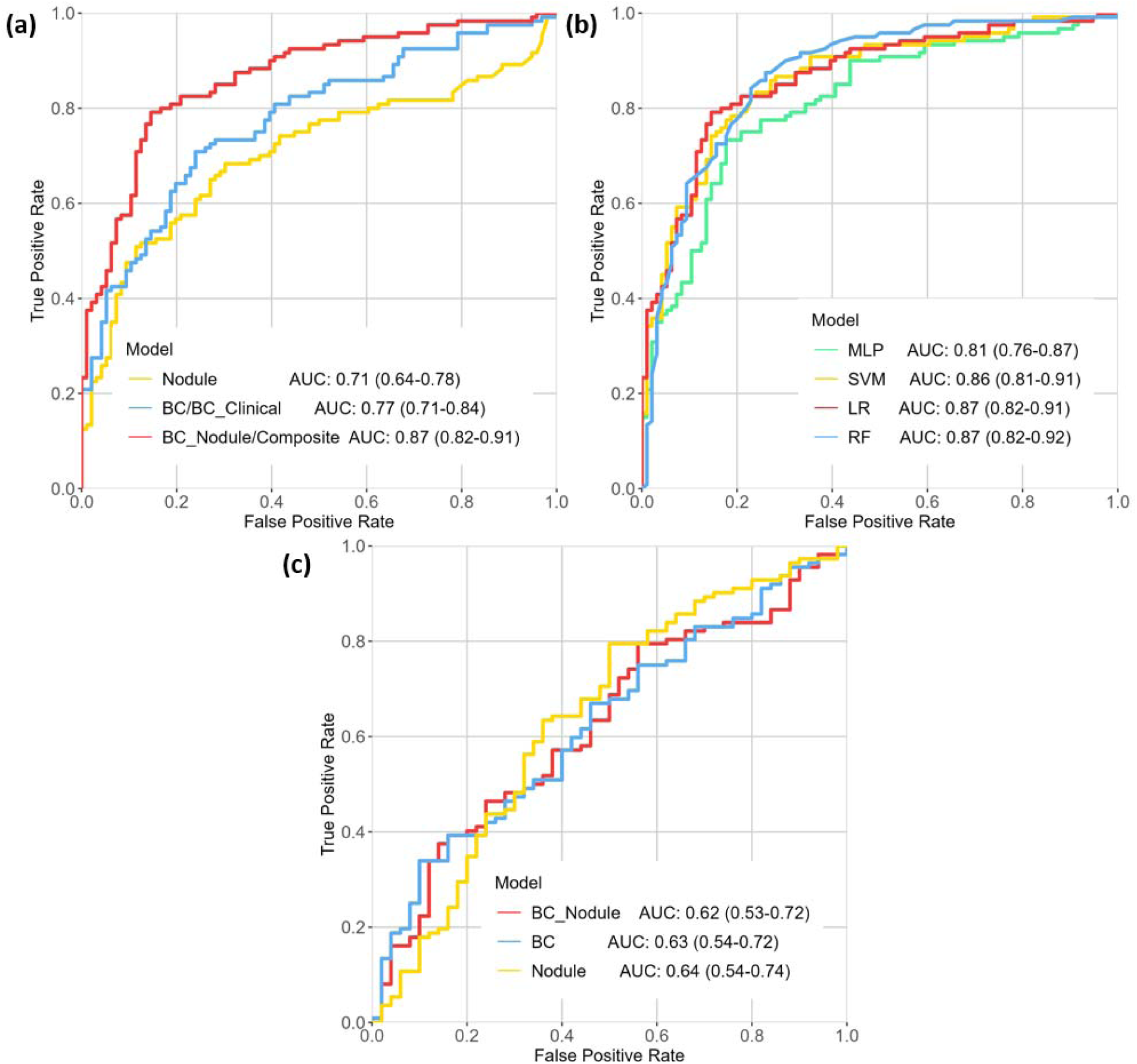
Cross-validated (10-fold) ROC-AUCs of (a) logistic regression integrative models (based on body composition and nodule features) using the internal dataset; (b) the best-performing *BC + Nodule (Composite)* model with different ML approaches; (c) integrative models on the external dataset.

**Table 3.** Multivariate logistic regression results of the integrative prediction models to discriminate benign and malignant nodules.

| Model | Variables included |  | Coefficient | P-value |
| --- | --- | --- | --- | --- |
| *Nodule* | Solidness |  | 0.60 | 0.037<sup>*</sup> |
|  | Irregularity |  | 0.88 | 0.000<sup>***</sup> |
|  | Calcification volume |  | -0.32 | 0.045<sup>*</sup> |
| *BC*<br>(*BC* + *Clinical*) | *Clinical* | none | / | / |
|  | *BC* | **IMAT density** | -0.47 | 0.023<sup>*</sup> |
|  |  | **SM density** | -1.49 | 0.000<sup>***</sup> |
|  |  | VAT density | -0.50 | 0.029<sup>*</sup> |
|  |  | **IMAT mass** | -0.84 | 0.000<sup>**</sup> |
|  |  | SAT mass | -0.64 | 0.003<sup>*</sup> |
| *BC + Nodule*<br>(*Composite*) | *Clinical* | none | / | / |
|  | *BC* | **IMAT density** | -0.79 | 0.001<sup>*</sup> |
|  |  | **SM density** | -2.18 | 0.000<sup>***</sup> |
|  |  | **IMAT mass** | -1.32 | 0.000<sup>**</sup> |
|  |  | SAT mass | -0.49 | 0.042<sup>*</sup> |
|  |  | **SM mass** | 0.77 | 0.003<sup>*</sup> |
|  | *Nodule* | Irregularity | 1.29 | 0.000<sup>***</sup> |
|  |  | Mean intensity | -0.79 | 0.000<sup>**</sup> |

*BC* — *Body Composition* model; *Composite* — *BC* + *Clinical* + *Nodule* model.
*Clinical* model is not presented due to its low performance.
After feature selection, *Clinical* features were not included in any models; *BC* and *BC* + *Clinical* model, *BC* + *Nodule* and *Composite* model retained the same features.
Coefficient and p-value from multivariate logistic regression model on standardized data.
<sup>\*</sup>, <sup>\*\*</sup>, <sup>\*\*\*</sup>, indicate p-value < 0.05, < 0.001, < 0.0001.
VAT: visceral adipose tissue, SAT: subcutaneous adipose tissue, IMAT: intermuscular adipose tissue, SM: skeletal muscle.

The individual *Nodule* model did not exhibit a significant performance difference compared to *BC* model (DeLong test: p = 0.151). Additionally, combining body composition features and nodule features, the *BC + Nodule* model demonstrated superior performance compared to the *Nodule* model alone (Likelihood ratio test: p < 0.0001).

### 3.2. Feature importance

The *BC + Nodule*/*Composite* model constructed using LR and RF methods achieved the highest performance, with an AUC of 0.87 (95% CI: 0.82 – 0.92), closely followed by SVM method with an AUC of 0.86 (95% CI: 0.81 – 0.91), and MLP method with an AUC of 0.81 (95% CI: 0.76 – 0.87) (Figure 3b). Feature importance analysis was conducted based on these four well-performing ML methods.

Among all body composition features, *SM density* was identified as the most important indicator and retained its significance even with the inclusion of *Nodule* features (Supplementary Table 6). Another crucial body composition feature is *IMAT* (Intermuscular adipose tissue), including both its density and mass. It is noteworthy that feature selection methods were not applied to the BC (body composition) feature importance analysis.

### 3.3. Causal analysis of body composition feature and lung nodule malignancy

Three features — body composition *SM density*, nodule features *Irregularity* and *Mean Intensity* — were directly linked to ‘malignancy’ (Figure 4). These variables demonstrated statistical significance in both univariate and multivariate logistic regression models and were highlighted as key indicators of nodule malignancy in feature importance analysis.

**Figure 4.**
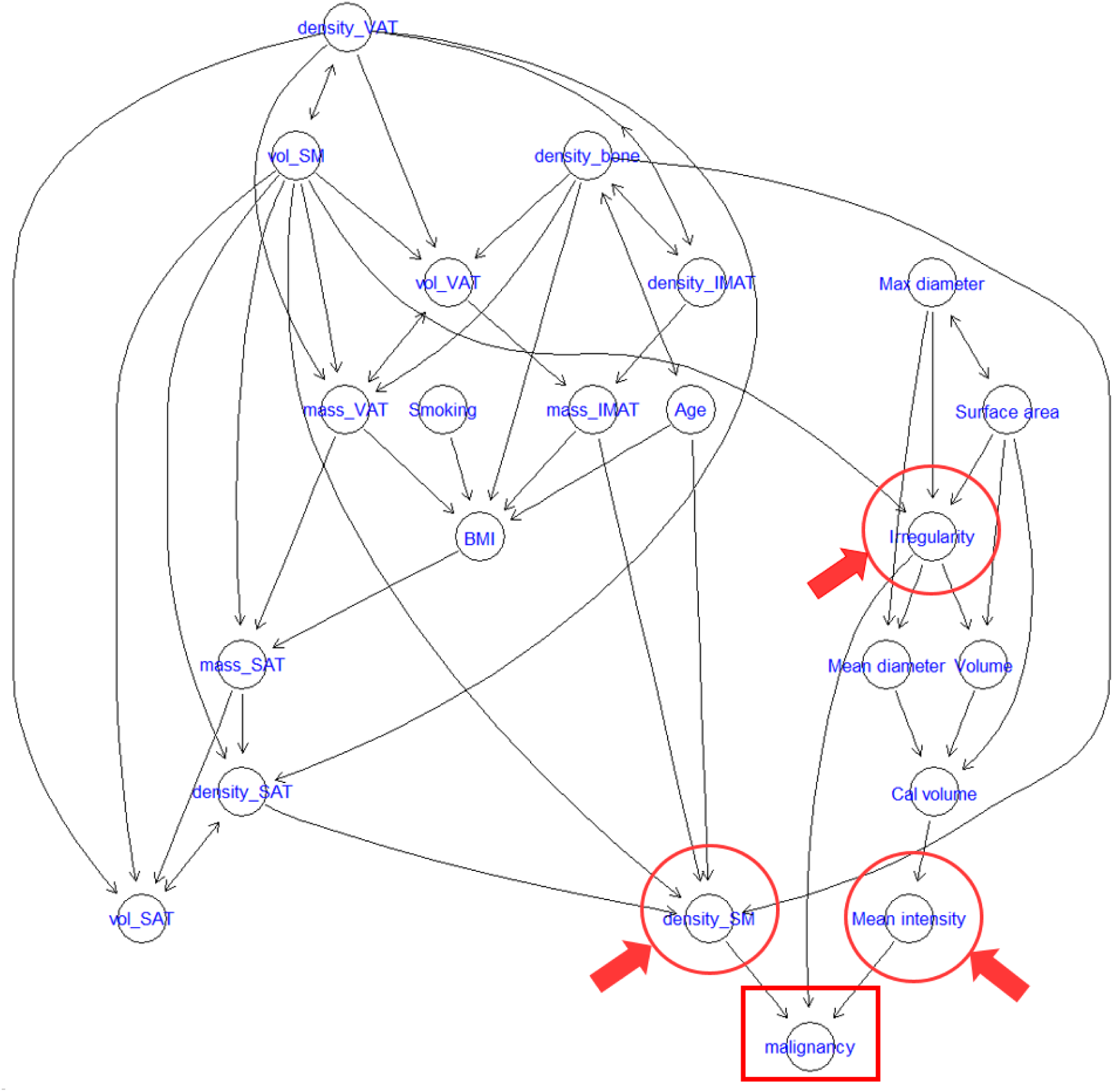
Graph-based causal relationship modeling among the variables. Only features with paths leading to the node ‘malignancy’ were retained in the graph. Two nodule features and body composition feature skeletal muscle density were linked to ‘malignancy’.

### 3.4. Body composition features and nodule changes

Nodule growth metrics were evaluated by identifying nodule features with significant changes between the baseline and the last LDCT scan date (Supplementary Table 7). These features were then used to analyze body composition changes. The correlations between changes in body composition and the identified nodule features are demonstrated in Supplementary Figure 4 for males and Supplementary Figure 5 for females. Body composition indicators related to nodule growth were identified for both genders (Table 4). Specifically, *IMAT density* and *SM density* were found to be important indicators of nodule growth in both males and females. *Bone* features were indicators of nodule growth in males. *SAT* features were identified as indicators in females.

**Table 4.** Identified nodule growth indicators.

| **Gender** | **Nodule** | **Body Tissue Composition** |
| --- | --- | --- |
| **Male** | Irregularity | Bone mass (-), Bone volume (-) |
|  | Mean diameter, Mean diameter (Solid) | **IMAT mass (+), IMAT density (+)** |
|  | Mean intensity | **SM density (-)** |
| **Female** | Irregularity, Surface area, Max diameter | **IMAT density (+)** |
|  | Mean diameter | SAT mass (+), SAT volume (+) |
|  | Mean intensity | **SM density (+)** |

A sign (+)/ (-) indicates positive/negative correlations between *Body Composition* and *Nodule* features.
SAT: subcutaneous adipose tissue, IMAT: intermuscular adipose tissue, SM: skeletal muscle.

Comparing the body composition changes from baseline to the last LDCT scans between malignant and benign groups (Supplementary Table 8), *IMAT* and *SM* features showed significant differences between the two groups in females. In males, all 3 *IMAT* features, *VAT density,* and *Bone density* were significantly different.

### 3.5. External validation

VUMC validation dataset included 112 (69.14%) malignant and 50 benign nodules, showing a higher malignancy rate than the 55.56% observed in the training dataset. Patients with malignant nodules in the external dataset were older, with an average age of 69.84 (± 8.41) years, compared to 63.20 (± 6.38) years in the training dataset. The average age of patients with benign nodules was similar across both datasets. While smoking status in the external dataset did not show a significant association with nodule malignancy, the number of cigarettes smoked per year showed a significant association.

Standardizing the external dataset using the same scales as the training dataset, we validated the trained baseline multivariate LR prediction models (Table 3) — including the individual *BC* model, *Nodule* model, and integrative *BC + Nodule* model — on the external dataset. All three models achieved very similar discrimination performance with AUCs of 0.64 (95% CI: 0.54 – 0.74), 0.63 (95% CI: 0.54 – 0.72), and 0.62 (95% CI: 0.53 – 0.72), respectively (Figure 3c). The performance of the three models was not significantly different (p > 0.05).

## 4. Discussion

A comprehensive study was conducted to investigate the associations between nodule malignancy, growth, and various image-based body composition features. Several critical body composition features were found to be associated with nodule malignancy and growth. Our main findings were validated on an external VUMC dataset and showed a lower but reasonable performance. To our knowledge, this is the first study to explore the potential relationships between body composition and nodule malignancy and growth using radiomic data, with a particular emphasis on gender differences.

We employed a robust feature selection procedure that combined LASSO regularization with iterative VIF selection and backward stepwise selection. This approach enhances predictive accuracy and mitigates overfitting issues, thereby improving model explainability. We validated this feature selection method through comprehensive feature importance analyses of body composition features without a pre-selection procedure (Supplementary Table 6). This strategy ensures the reliability of our procedure for feature selection. Additionally, we utilized a causal graph to analyze and visualize the causal relationships between body composition features and malignancy.

The CT-derived body composition features were consistently and significantly associated with nodule malignancy in all analyses. Most notably, the loss of *SM* (Skeletal muscle) and *IMAT* (Intramuscular adipose tissue) were highlighted. Muscle wasting (sarcopenia) is recognized as a prominent phenotype in lung cancer patients [12, 27]. However, previous research has either characterized muscle loss by comparing lung cancer and non-lung cancer groups or studied its association with the overall survival of lung cancer patients. Our study suggests that muscle wasting is crucial for indicating a lung nodule’s malignancy on LDCT scans.

Clinical and CT-derived nodule features are widely studied and acknowledged biomarkers for nodule malignancy [7]. Key clinical features, such as age, sex, smoking history, and nodule features, especially nodule size, are commonly utilized in lung cancer prediction models [7]. However, in our analysis, only *Emphysema* was found to be significant univariately (Table 1). None of the clinical features were retained in the integrative models after feature selection (Table 3). Potential reasons for this could be: (1) the models retained body tissue features over age because the aging process is often paralleled by decreases in muscle and increases in fat mass [28]; (2) gender, age, BMI, and smoking history was balanced between the malignant and benign groups in our dataset, which could reduce the bias and effects of these features on malignancy assessment. Additionally, previous studies [29, 30] have suggested that the existence of emphysema could help predict lung cancer risk among smokers. In our study, after the inclusion of body tissue features (Table 3), emphysema was excluded because tissue features were more significant for nodule classification. Multiple nodule features were found to be associated with nodule malignancy both univariately and multivariately. The causal graph (Figure 4) underscored the significance of skeletal muscle density, as nodule features serve as direct indicators of nodule malignancy. The graph links two nodule features, *Irregularity* and *Mean Intensity*, along with *SM density*, directly to node ‘malignancy’.

The performance of the *Nodule* and *BC* models did not show a significant performance difference, whereas the *BC + Nodule* model demonstrated superior performance over the *Nodule* model. These findings suggest: (1) body composition can serve as a novel biomarker of nodule malignancy assessment; and (2) combining body composition with nodule features could enhance nodule malignancy assessment. The first finding was validated in an external dataset. Despite the *BC* model not achieving high performance, its performance was comparable to that of the *Nodule* model, indicating the classification ability of body composition features based on LDCT image data. The relatively low performance in the VUMC dataset could be attributed to the variations in image protocols and patient characteristics between the two datasets. Specifically, the PLuSS training dataset had more balanced clinical characteristics compared to the VUMC validation dataset (Table 1, Supplementary Table 1).

Body composition differs significantly between males and females, prompting the need for gender-specific analyses in lung cancer studies [12, 14, 15]. Our study similarly accounted for these gender differences to ensure a more accurate assessment. Specifically, in the analysis of malignancy, *SM density* remained a malignancy indicator for both males and females, while *Bone* features were significant only for males and *VAT*, *IMAT* features only for females. In the analysis of nodule growth indicators (Table 4, Supplementary Table 8), the significance of *IMAT* and *SM* features was highlighted again for both genders. Additionally, *Bone* features (bone loss) were identified as an indicator of nodule growth in males, given that males tend to have higher bone density than females [31]. We observed that *SM density* (Skeletal muscle density) change was negatively correlated with nodule *Mean intensity* in males but positively correlated in females. Two possible explanations: (1) different body composition in males and females, such as denser muscles with more intramuscular fat in males than in females, might influence these correlations; and (2) although nodule *Mean intensity* can provide important clues about the likelihood of malignancy (Table 2) and nodule growth (Supplementary Table 7), it is important to note that a sole change in nodule density does not necessarily indicate malignant nodule growth. For example, a nodule density increase can indicate either calcification [32] (a benign process) or malignant nodules growing from partly solid to more solid [33], while a nodule density decrease can indicate either inflammation of benign nodules [34] or cavitation of malignant nodules [35] as they grow. When considering nodule malignancy, it is necessary to combine nodule density with other features, such as nodule size and irregularity (Table 3). Overall, nodule density change is still an important indicator of nodule growth.

The primary limitation of this study is the inability to validate the relationship between body composition and nodule growth in the external dataset. This limitation arises from the lack of follow-up CT scans in the external dataset. Additionally, although our models revealed a potential relationship between body composition and nodule malignancy on the external dataset, the prediction performance was relatively lower compared to the internal dataset. This discrepancy may be due to variations in CT acquisition protocol, participant enrollment criteria, and population characteristics. Nonetheless, our results indicate a strong connection between body composition and nodule malignancy, as well as their changes over time.

## 5. Conclusion

We performed a comprehensive investigation of the body composition as a potential biomarker for assessing pulmonary nodule malignancy and growth. We found that body composition, especially *skeletal muscle density* and *intermuscular adipose tissue density*, were critical indicators for discriminating malignant nodules from benign ones and for evaluating the nodule growth in both males and females. These findings can help significantly improve lung cancer screening by improving the assessment of the malignancy of screen-detected lung nodules. Given the high prevalence of false positive nodule detections during lung cancer screening, our findings have significant clinical implications. Additionally, our findings can help evaluate nodule progression or stability, leading to more accurate prognoses and personalized management strategies.

## Data Availability

All data produced in the present study are available upon reasonable request to the authors

## Abbreviations

AI: artificial intelligence
BC: body composition
CI: confidence interval
LDCT: low-dose computed tomography
LASSO: least absolute shrinkage and selection operator
LR: logistic regression
PI: permutation importance
ROC-AUC: the area under the receiver operating characteristic curve
RF: random forest
SVM: support vector machine

## References

1. Siegel, R.L., A.N. Giaquinto, and A. Jemal, Cancer statistics, 2024. CA: a cancer journal for clinicians, 2024. 74(1): p. 12–49.

2. Wender, R., et al., American Cancer Society lung cancer screening guidelines. CA: a cancer journal for clinicians, 2013. 63(2): p. 106–117.

3. Reduced Lung-Cancer Mortality with Low-Dose Computed Tomographic Screening. New England Journal of Medicine, 2011. 365(5): p. 395–409.

4. Chudgar, N.P., et al., Results of the National Lung Cancer Screening Trial. Thoracic Surgery Clinics, 2015. 25(2): p. 145–153.

5. Marques, S., et al., A multi-task CNN approach for lung nodule malignancy classification and characterization. Expert Systems with Applications, 2021. 184: p. 115469.

6. Chen, H., et al., Intra-and Inter-Reader Variations in Lung Nodule Measurements: Influences of Nodule Size, Location, and Observers. Diagnostics, 2022. 12(10): p. 2319.

7. Zhang, K., et al., Comprehensive analysis of clinical logistic and machine learning-based models for the evaluation of pulmonary nodules. JTO Clinical and Research Reports, 2022. 3(4): p. 100299.

8. Way, T.W., et al., Computer aided diagnosis of pulmonary nodules on CT scans: Improvement of classification performance with nodule surface features. Medical Physics, 2009. 36(7): p. 3086–3098.

9. Armato III, S.G., et al., LUNGx Challenge for computerized lung nodule classification. Journal of Medical Imaging, 2016. 3(4): p. 044506–044506.

10. Vachani, A., et al., Factors that influence physician decision making for indeterminate pulmonary nodules. Annals of the American Thoracic Society, 2014. 11(10): p. 1586–1591.

11. Nibali, A., Z. He, and D. Wollersheim, Pulmonary nodule classification with deep residual networks. International Journal of Computer Assisted Radiology and Surgery, 2017. 12(10): p. 1799–1808.

12. Baracos, V.E., et al., Body composition in patients with non− small cell lung cancer: a contemporary view of cancer cachexia with the use of computed tomography image analysis. The American journal of clinical nutrition, 2010. 91(4): p. 1133S–1137S.

13. Baldessari, C., et al., Body composition and inflammation impact in non-small-cell lung cancer patients treated by first-line immunotherapy. Immunotherapy, 2021. 13(18): p. 1501–1519.

14. Rizzo, S., et al., CT-derived body composition values and complications after pneumonectomy in lung cancer patients: time for a sex-related analysis? Frontiers in Oncology, 2022. 12: p. 826058.

15. Martin, L., et al., Cancer cachexia in the age of obesity: skeletal muscle depletion is a powerful prognostic factor, independent of body mass index. Journal of clinical oncology, 2013. 31(12): p. 1539–1547.

16. Sjøblom, B., et al., Skeletal muscle radiodensity is prognostic for survival in patients with advanced non-small cell lung cancer. Clinical Nutrition, 2016. 35(6): p. 1386–1393.

17. Degens, J.H., et al., The prognostic value of early onset, CT derived loss of muscle and adipose tissue during chemotherapy in metastatic non-small cell lung cancer. Lung Cancer, 2019. 133: p. 130–135.

18. Lee, H.Y. and K.S. Lee, Ground-glass opacity nodules: histopathology, imaging evaluation, and clinical implications. Journal of thoracic imaging, 2011. 26(2): p. 106–118.

19. Wilson, D.O., et al., The Pittsburgh Lung Screening Study (PLuSS) outcomes within 3 years of a first computed tomography scan. American journal of respiratory and critical care medicine, 2008. 178(9): p. 956–961.

20. Xiao, D., et al., Assessing the Transportability of Radiomic Models for Lung Cancer Diagnosis. Translational Lung Cancer Research, 2024 (In Press).

21. Pu, L., et al., Automated segmentation of five different body tissues on computed tomography using deep learning. Medical Physics, 2023. 50(1): p. 178–191.

22. Pu, L., et al., Estimating 3-D whole-body composition from a chest CT scan. Med Phys, 2022. 49(11): p. 7108–7117.

23. Ashraf, S.F., et al., Predicting benign, preinvasive, and invasive lung nodules on computed tomography scans using machine learning. J Thorac Cardiovasc Surg, 2022. 163(4): p. 1496–1505 e10.

24. Pu, J., et al., Shape “break-and-repair” strategy and its application to automated medical image segmentation. IEEE transactions on visualization and computer graphics, 2010. 17(1): p. 115–124.

25. Lundberg, S.M. and S.-I. Lee, A unified approach to interpreting model predictions. Advances in neural information processing systems, 2017. 30.

26. Ren, S., et al., Graphical modeling of causal factors associated with the postoperative survival of esophageal cancer subjects. Medical Physics, 2024. 51(3): p. 1997–2006.

27. Al-Sawaf, O., et al., Body composition and lung cancer-associated cachexia in TRACERx. Nature medicine, 2023. 29(4): p. 846–858.

28. Larsson, L., et al., Sarcopenia: aging-related loss of muscle mass and function. Physiological reviews, 2019. 99(1): p. 427–511.

29. Li, Y., et al., Effect of emphysema on lung cancer risk in smokers: a computed tomography–based assessment. Cancer prevention research, 2011. 4(1): p. 43–50.

30. Mouronte-Roibas, C., et al., COPD, emphysema and the onset of lung cancer. A systematic review. Cancer letters, 2016. 382(2): p. 240–244.

31. Nieves, J.W., et al., Males have larger skeletal size and bone mass than females, despite comparable body size. Journal of Bone and Mineral Research, 2005. 20(3): p. 529–535.

32. Khan, A.N., et al., The calcified lung nodule: What does it mean? Annals of thoracic medicine, 2010. 5(2): p. 67–79.

33. Xu, D.M., et al., Role of baseline nodule density and changes in density and nodule features in the discrimination between benign and malignant solid indeterminate pulmonary nodules. European Journal of Radiology, 2009. 70(3): p. 492–498.

34. Kamiya, A., et al., Kurtosis and skewness assessments of solid lung nodule density histograms: differentiating malignant from benign nodules on CT. Japanese journal of radiology, 2014. 32: p. 14–21.

35. Chaudhry, A., et al., Characteristic CT findings after percutaneous cryoablation treatment of malignant lung nodules. Medicine, 2015. 94(42): p. e1672.

