## Supplemental Material for "CT-derived Body Composition Associated with Pulmonary Nodule Malignancy and Growth"

**Supplementary Materials:**

**S. Table 1.** Subject demographic and clinical characteristics of the XXX validation dataset (n=162).

| Variable | | Malignant (n=112) | Benign (n=50) | Coefficient (95% CI) | P-value |
| --- | --- | --- | --- | --- | --- |
| Age (year) | | 69.84 (8.41) | 62.88 (10.07) | 0.09 (0.04, 0.13) | 0.000^***^ |
| Gender |  |  |  |  |  |
|  | Male | 67 | 27 | 0.24 (-0.43, 0.91) | 0.488 |
|  | Female | 45 | 23 |  |  |
| BMI | | 26.98 (7.08) | 29.59 (7.83) | -0.05 (-0.10, -0.00) | 0.041^*^ |
| Smoking Status |  |  |  |  |  |
|  | Current | 104 | 42 | 0.91 (-0.14, 1.95) | 0.089 |
|  | Former | 8 | 8 |  |  |
| Pack (year) | | 49.13 (32.67) | 30.1 (23.03) | 0.03 (0.01, 0.04) | 0.000^**^ |
| Mean (Standard deviation (SD)) and Count for continuous and categorical variables respectively. | | | | | |
| Coefficient and p-value from univariate logistic regression model on unstandardized data. | | | | | |
| ^*^, ^**^, ^***^, indicate p-value < 0.05, < 0.001, < 0.0001. | | | | | |
| Values of 0.00 are non-zero but are truncated due to rounding. | | | | | |

**S. Table 2.** Summary statistics of CT-derived body composition in lung cancer and non-cancer groups and their univariate logistic regression results /Male (n=113).

| Variable | Malignant (n=61) | Benign (n=52) | Coefficient (95% CI) | P-value |
| --- | --- | --- | --- | --- |
| VAT volume (L) | 1.57 (0.86) | 1.40 (0.61) | 0.29 (-0.21, 0.80) | 0.256 |
| VAT density (HU) | -91.15 (5.93) | -90.08 (4.86) | -0.04 (-0.11, 0.03) | 0.300 |
| VAT mass (kg) | 1.62 (0.88) | 1.45 (0.63) | 0.28 (-0.21, 0.78) | 0.262 |
| SAT volume (L) | 3.49 (1.40) | 4.20 (1.84) | -0.28 (-0.53, -0.03) | **0.028^*^** |
| SAT density (HU) | -85.37 (10.84) | -85.10 (8.60) | -0.00 (-0.04, 0.04) | 0.884 |
| SAT mass (kg) | 3.63 (1.44) | 4.37 (1.88) | -0.28 (-0.52, -0.03) | **0.025^*^** |
| IMAT volume (L) | 0.53 (0.29) | 0.55 (0.26) | -0.16 (-1.53, 1.20) | 0.813 |
| IMAT density (HU) | -91.53 (14.46) | -88.06 (15.37) | -0.02 (-0.04, 0.01) | 0.220 |
| IMAT mass (kg) | 0.55 (0.29) | 0.57 (0.26) | -0.21 (-1.56, 1.14) | 0.761 |
| SM volume (L) | 5.08 (0.77) | 5.31 (0.75) | -0.40 (-0.91, 0.10) | 0.116 |
| SM density (HU) | 30.82 (7.17) | 35.37 (6.09) | -0.10 (-0.16, -0.04) | **0.001^**^** |
| SM mass (kg) | 5.94 (0.89) | 6.24 (0.90) | -0.38 (-0.81, 0.05) | 0.085 |
| Bone volume (L) | 1.86 (0.24) | 1.75 (0.23) | 1.98 (0.30, 3.66) | **0.021^*^** |
| Bone density (HU) | 267.15 (33.49) | 283.42 (46.34) | -0.01 (-0.02, -0.00) | **0.038^*^** |
| Bone mass (kg) | 2.65 (0.35) | 2.53 (0.35) | 1.04 (-0.07, 2.15) | 0.067 |
| Mean (Standard deviation (SD)) for continuous variables. | | | | |
| Coefficient and p-value from univariate logistic regression model on unstandardized data. | | | | |
| ^*^, ^**^, indicate p-value < 0.05, < 0.001. | | | | |
| Values of 0.00 are non-zero but are truncated due to rounding. | | | | |
| VAT: visceral adipose tissue, SAT: subcutaneous adipose tissue, IMAT: intermuscular adipose tissue, | | | | |
| SM: skeletal muscle. | | | | |

**S. Table 3.** Summary statistics of CT-derived body composition in lung cancer and non-cancer groups and their univariate logistic regression results /Female (n=103).

| Variable | Malignant (n=59) | Benign (n=44) | Coefficient (95% CI) | P-value |
| --- | --- | --- | --- | --- |
| VAT volume (L) | 0.70 (0.45) | 0.63 (0.44) | 0.42 (-0.50, 1.34) | 0.374 |
| VAT density (HU) | -89.33 (5.9) | -86.61 (5.46) | -0.09 (-0.16, -0.01) | **0.023^*^** |
| VAT mass (kg) | 0.73 (0.46) | 0.65 (0.45) | 0.40 (-0.49, 1.29) | 0.379 |
| SAT volume (L) | 4.58 (1.73) | 4.12 (1.74) | 0.16 (-0.08, 0.40) | 0.190 |
| SAT density (HU) | -93.60 (7.05) | -89.55 (8.16) | -0.07 (-0.13, -0.02) | **0.012^*^** |
| SAT mass (kg) | 4.72 (1.76) | 4.26 (1.78) | 0.15 (-0.08, 0.39) | 0.198 |
| IMAT volume (L) | 0.38 (0.16) | 0.35 (0.16) | 1.20 (-1.38, 3.77) | 0.363 |
| IMAT density (HU) | -88.67 (9.11) | -84.64 (9.66) | -0.05 (-0.10, -0.00) | **0.037^*^** |
| IMAT mass (kg) | 0.39 (0.16) | 0.37 (0.16) | 1.11 (-1.39, 3.61) | 0.383 |
| SM volume (L) | 3.25 (0.61) | 3.34 (0.42) | -0.31 (-1.05, 0.44) | 0.420 |
| SM density (HU) | 27.13 (6.79) | 32.98 (5.82) | -0.14 (-0.21, -0.07) | **0.000^**^** |
| SM mass (kg) | 3.79 (0.71) | 3.91 (0.49) | -0.32 (-0.96, 0.33) | 0.335 |
| Bone volume (L) | 1.26 (0.19) | 1.21 (0.12) | 2.41 (-0.40, 5.21) | 0.092 |
| Bone density (HU) | 278.12 (37.16) | 291.68 (44.63) | -0.01 (-0.02, 0.00) | 0.097 |
| Bone mass (kg) | 1.82 (0.27) | 1.76 (0.19) | 1.14 (-0.64, 2.92) | 0.210 |
| Mean (Standard deviation (SD)) for continuous variables. | | | | |
| Coefficient and p-value from univariate logistic regression model on unstandardized data. | | | | |
| ^*^, ^**^, indicate p-value < 0.05, < 0.001. | | | | |
| Values of 0.00 are non-zero but are truncated due to rounding. | | | | |
| VAT: visceral adipose tissue, SAT: subcutaneous adipose tissue, IMAT: intermuscular adipose tissue, | | | | |
| SM: skeletal muscle. | | | | |

**S. Table 4.** Summary statistics of CT-derived nodule features in lung cancer and non-cancer groups and their univariate logistic regression results /Male (n=113).

| Variable | Malignant (n=61) | Benign (n=52) | Coefficient (95% CI) | P-value |
| --- | --- | --- | --- | --- |
| Volume (ml) | 0.57 (1.15) | 0.38 (1.05) | 0.18 (-0.22, 0.58) | 0.377 |
| Mean intensity (HU) | -396.49 (170.25) | -287.43 (181.93) | -0.00 (-0.01, -0.00) | **0.003^*^** |
| Surface area (cm^2^) | 3.30 (4.28) | 2.17 (3.48) | 0.09 (-0.03, 0.20) | 0.152 |
| Max diameter (mm) | 10.17 (5.63) | 7.77 (4.70) | 0.01 (-0.00, 0.02) | **0.023^*^** |
| Mean diameter (mm) | 7.49 (4.15) | 6.29 (3.69) | 0.08 (-0.02, 0.19) | **0.048^*^** |
| Mean diameter (Solid) (mm) | 5.22 (4.04) | 4.98 (3.71) | 0.02 (-0.08, 0.11) | 0.743 |
| Solidness | 0.07 (0.15) | 0.01 (0.02) | 18.04 (3.91, 32.18) | **0.012^*^** |
| Calcification volume (mm^3^) | 46.26 (233.77) | 28.91 (87.44) | 0.00 (-0.00, 0.00) | 0.623 |
| Irregularity (%) | 0.33 (0.28) | 0.14 (0.17) | 4.03 (1.89, 6.17) | **0.000^**^** |
| Mean (Standard deviation (SD)) for continuous variables. | | | | |
| Coefficient and p-value from univariate logistic regression model on unstandardized data. | | | | |
| ^*^, ^**^, indicate p-value < 0.05, < 0.001. | | | | |
| Values of 0.00 are non-zero but are truncated due to rounding. | | | | |

**S. Table 5.** Summary statistics of CT-derived nodule features in lung cancer and non-cancer groups and their univariate logistic regression results /Female (n=103).

| Variable | Malignant (n=59) | Benign (n=44) | Coefficient (95% CI) | P-value |
| --- | --- | --- | --- | --- |
| Volume (ml) | 0.51 (1.20) | 0.37 (1.43) | 0.09 (-0.25, 0.43) | 0.594 |
| Mean intensity (HU) | -350.17 (203.57) | -230.67 (310.48) | -0.00 (-0.00, -0.00) | **0.026^*^** |
| Surface area (cm^2^) | 3.16 (4.25) | 2.19 (5.42) | 0.05 (-0.05, 0.16) | 0.335 |
| Max diameter (mm) | 9.99 (6.34) | 7.31 (4.92) | 0.02 (0.00, 0.03) | **0.034^*^** |
| Mean diameter (mm) | 7.36 (4.15) | 5.63 (3.03) | 0.16 (0.02, 0.29) | **0.028^*^** |
| Mean diameter (Solid) (mm) | 7.99 (20.46) | 4.21 (2.76) | 0.10 (-0.02, 0.23) | 0.115 |
| Solidness | 0.07 (0.12) | 0.02 (0.05) | 7.23 (1.05, 13.40) | **0.022^*^** |
| Calcification volume (mm^3^) | 42.80 (132.79) | 88.87 (409.08) | -0.00 (-0.00, 0.00) | 0.450 |
| Irregularity (%) | 0.36 (0.29) | 0.18 (0.19) | 3.23 (1.21, 5.25) | **0.002^*^** |
| Mean (Standard deviation (SD)) for continuous variables. | | | | |
| Coefficient and p-value from univariate logistic regression model on unstandardized data. | | | | |
| ^*^, indicates p-value < 0.05. | | | | |
| Values of 0.00 are non-zero but are truncated due to rounding. | | | | |

**S. Table 6.** Feature importance.

| **Model** | **Features** | **PI Score** | | | | **SHAP Score** | | | | **Average**  **Score** |
| --- | --- | --- | --- | --- | --- | --- | --- | --- | --- | --- |
|  |  | LR | SVM | RF | MLP | LR | SVM | RF | MLP |  |
| BC (3) ^*^ | **SM density** | 0.13 | 1.00 | 1.00 | 1.00 | 0.02 | 1.00 | 1.00 | 1.00 | 0.78 |
|  | **IMAT density** | 0.23 | 0.24 | 0.72 | 0.71 | 0.11 | 0.38 | 0.60 | 0.69 | 0.47 |
|  | SM mass | 1.00 | 0.10 | 0.25 | 0.39 | 1.00 | 0.05 | 0.03 | 0.23 | 0.44 |
| *BC + Nodule*  *(Composite)* | **SM density** | 1.00 | 1.00 | 0.92 | 1.00 | 1.00 | 1.00 | 1.00 | 1.00 | 0.98 |
|  | Irregularity | 0.64 | 0.63 | 1.00 | 0.87 | 0.19 | 0.36 | 0.76 | 0.35 | 0.78 |
|  | **IMAT mass** | 0.53 | 0.48 | 0.31 | 0.48 | 0.26 | 0.30 | 0.00 | 0.00 | 0.45 |
|  | Mean intensity | 0.31 | 0.26 | 0.33 | 0.52 | 0.02 | 0.10 | 0.33 | 0.11 | 0.36 |
|  | **IMAT density** | 0.25 | 0.17 | 0.52 | 0.37 | 0.05 | 0.08 | 0.53 | 0.22 | 0.33 |
|  | SAT mass | 0.08 | 0.00 | 0.14 | 0.27 | 0.00 | 0.00 | 0.37 | 0.23 | 0.12 |
|  | SM mass | 0.00 | 0.05 | 0.00 | 0.00 | 0.20 | 0.28 | 0.01 | 0.18 | 0.01 |
| ^*^ Only the top three body composition features were listed based on their average importance scores. | | | | | | | | | | |
| *BC + Nodule (Composite)* model — the best-performing multivariate prediction model. | | | | | | | | | | |
| SAT: subcutaneous adipose tissue, IMAT: intermuscular adipose tissue, SM: skeletal muscle, | | | | | | | | | | |
| PI: permutation importance, LR: logistic regression, SVM: support vector machine, | | | | | | | | | | |
| RF: random forest, MLP: multi-layer perceptron. | | | | | | | | | | |

**S. Table 7.** Nodule growth metrics and t-tests (n=216).

| **Variable** | **Baseline** | **Last follow-up** | **Absolute Change** | **P-value (T-test)** |
| --- | --- | --- | --- | --- |
| **Volume (ml)** | 0.47 (1.20) | 1.14 (2.30) | 0.68 (2.01) | 0.000^***^ |
| **Mean intensity (HU)** | -323.59 (224.10) | -269.11 (205.82) | 54.48 (169.20) | 0.000^***^ |
| **Surface area (cm^2^)** | 2.76 (4.36) | 5.30 (6.92) | 2.54 (5.84) | 0.000^***^ |
| **Max diameter (mm)** | 8.96 (5.61) | 12.75 (7.75) | 3.79 (6.19) | 0.000^***^ |
| **Mean diameter (mm)** | 6.79 (3.88) | 9.59 (5.46) | 2.81 (4.37) | 0.000^***^ |
| **Mean diameter (Solid) (mm)** | 5.73 (11.24) | 7.69 (5.35) | 1.97 (11.81) | 0.015^*^ |
| Solidness | 0.05 (0.11) | 0.06 (0.11) | 0.01 (0.11) | 0.075 |
| Calcification volume (mm^3^) | 49.80 (235.70) | 60.75 (194.22) | 10.95 (227.60) | 0.480 |
| **Irregularity (%)** | 0.26 (0.26) | 0.36 (0.29) | 0.09 (0.25) | 0.000^***^ |
| Mean (Standard deviation (SD)) for continuous variables. | | | | |
| ^*^, ^**^, ^***^, indicate p-value < 0.05, < 0.001, < 0.0001. | | | | |

**S. Table 8.** Absolute change comparison of body composition features between malignant and benign groups in both males and females.

| **Variable** | **P-values (Male=113)** | **P-values (Female=103)** |
| --- | --- | --- |
| VAT volume_delta | 0.182 | 0.177 |
| VAT density_delta | **0.018^*^** | 0.180 |
| VAT mass_delta | 0.190 | 0.170 |
| SAT volume_delta | 0.367 | 0.066 |
| SAT_density_delta | 0.483 | 0.396 |
| SAT mass_delta | 0.387 | 0.066 |
| IMAT volume_delta | **0.009^*^** | 0.263 |
| IMAT density_delta | **0.008^*^** | **0.018^*^** |
| IMAT mass_delta | **0.011^*^** | 0.291 |
| SM volume_delta | 0.695 | **0.024^*^** |
| SM density_delta | 0.442 | 0.688 |
| SM mass_delta | 0.712 | **0.023^*^** |
| Bone volume_delta | 0.412 | 0.102 |
| Bone density_delta | **0.001^**^** | 0.918 |
| Bone mass_delta | 0.142 | 0.141 |
| Delta was computed using data from the baseline and last follow-up dates. | | |
| P-values from T-tests comparing malignant and benign groups. | | |
| ^*^, ^**^, indicate p-value < 0.05, < 0.001. | | |
| VAT: visceral adipose tissue, SAT: subcutaneous adipose tissue,  IMAT: intermuscular adipose tissue, SM: skeletal muscle. | | |

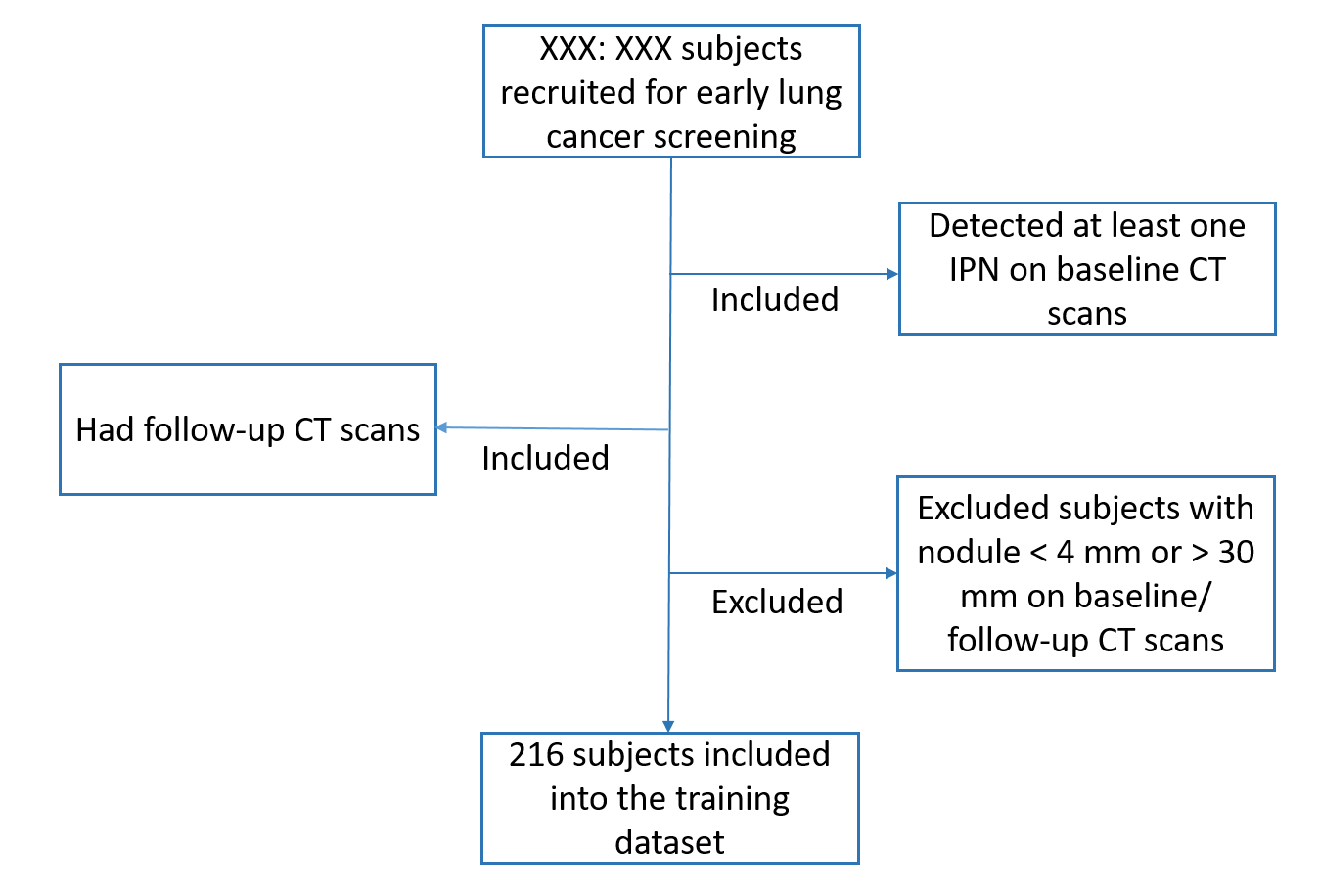

**S. Figure 1.** Subject inclusion and exclusion procedure for the training dataset. XXX.

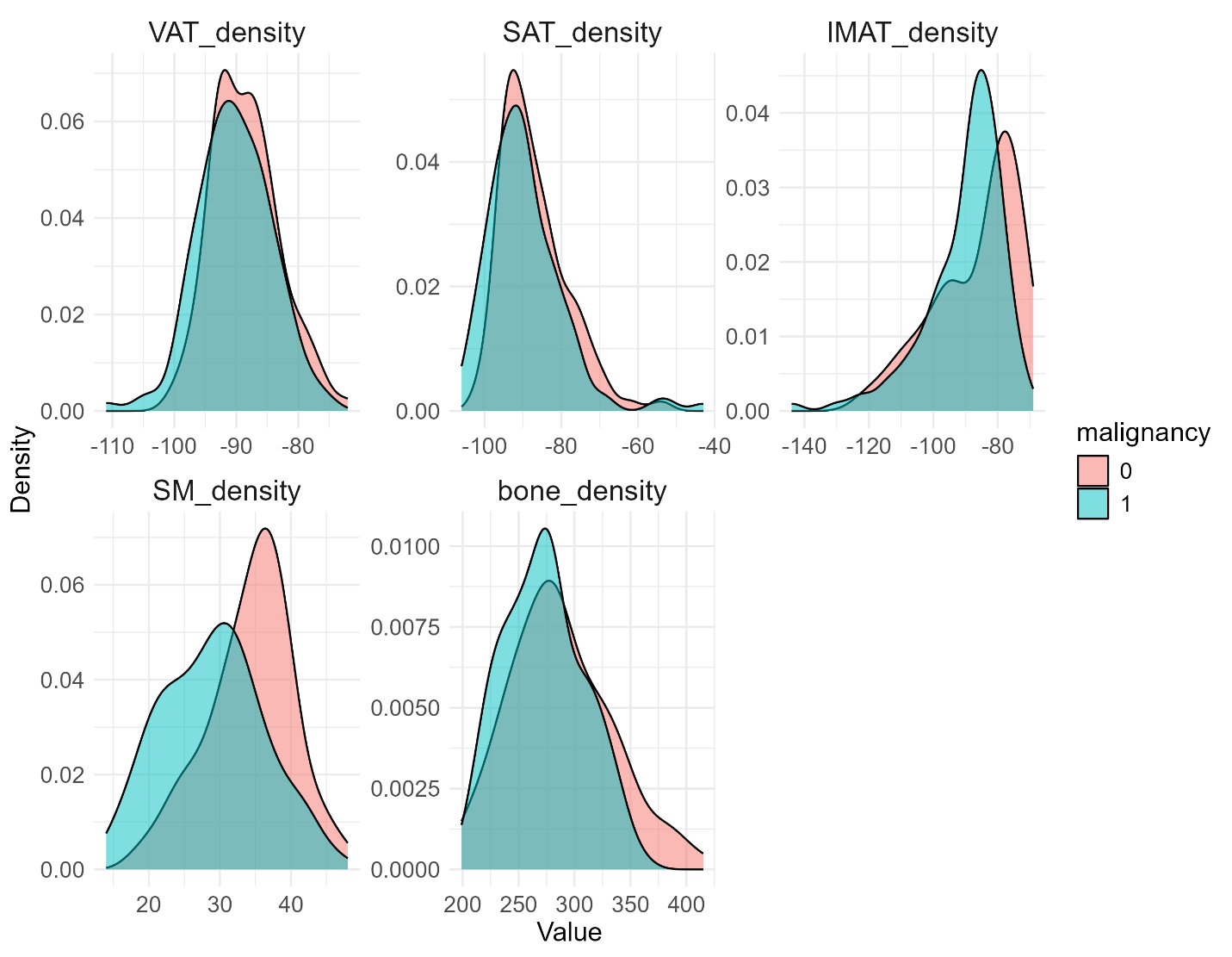

**S. Figure 2.** Distributions of five body tissue densities by malignancy

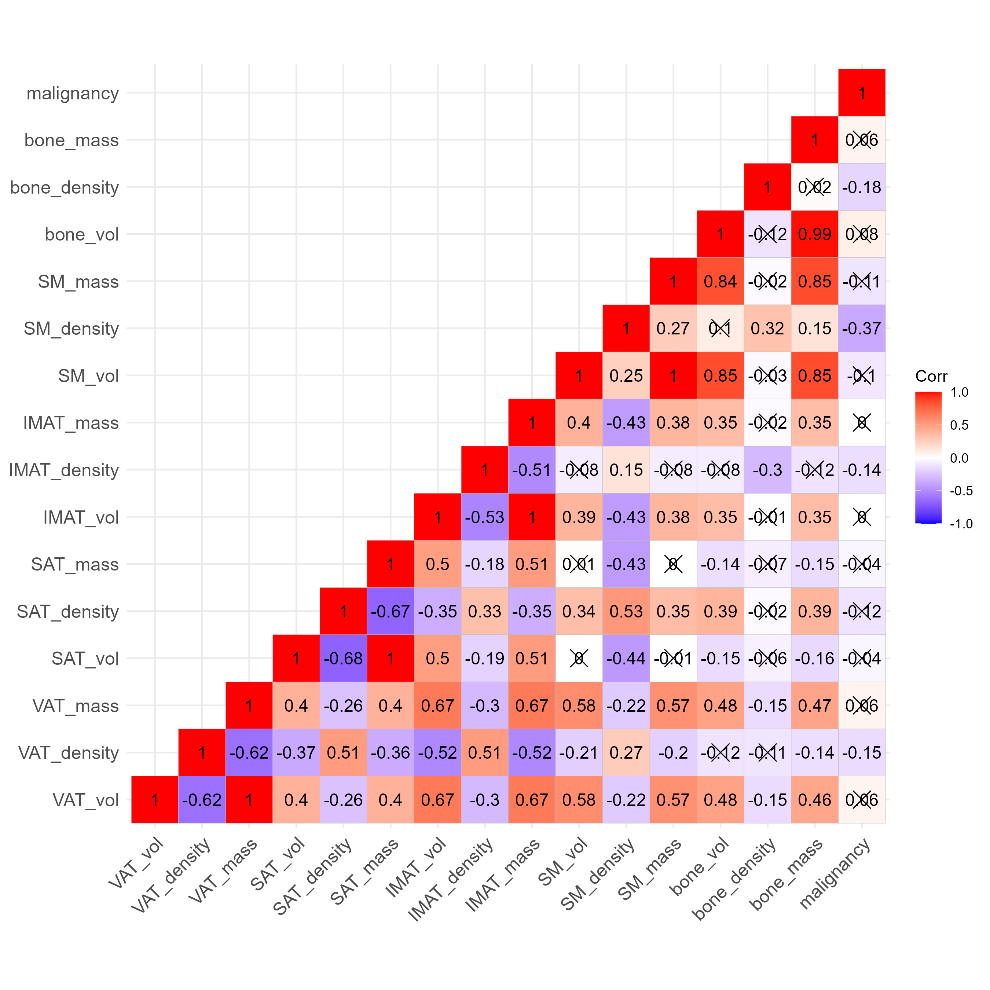

**S. Figure 3.** Correlation visualization between baseline body composition and malignancy (both genders). Correlations marked with a cross are statistically insignificant (p-value > 0.05).

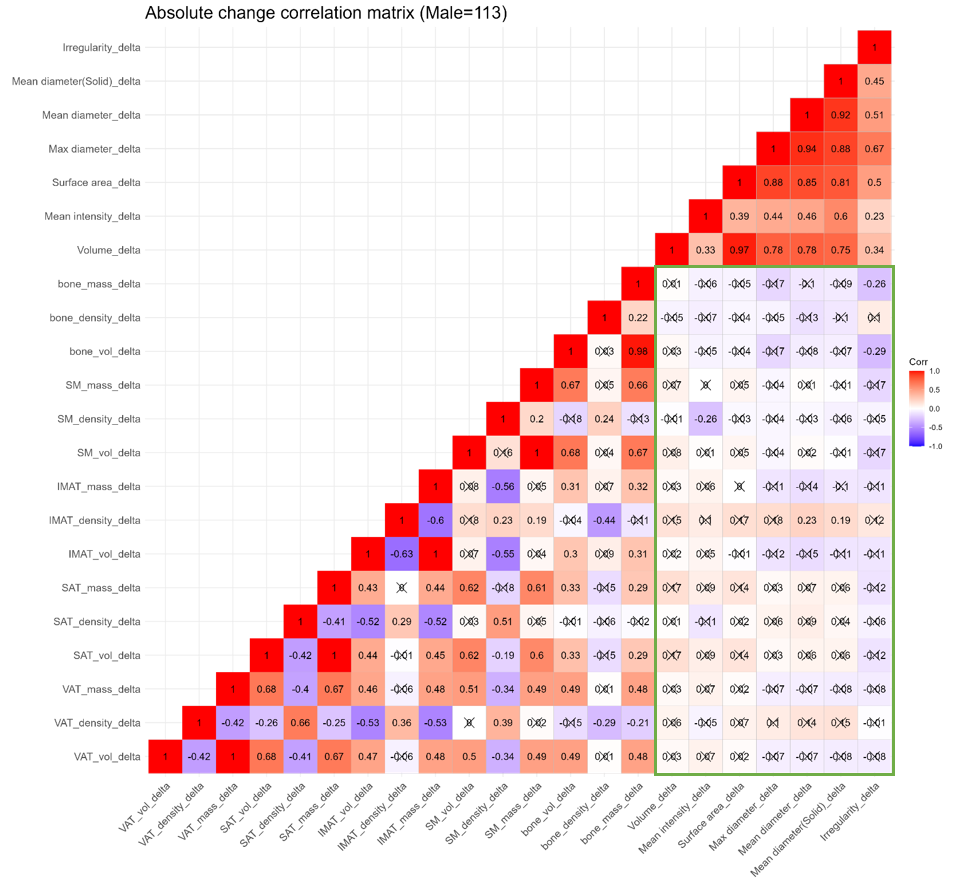

**S. Figure 4.** Correlation visualization between changes of body composition and nodule features (Male=113). Correlations marked with a cross are statistically insignificant (p-value > 0.05). The correlations between nodule and body composition features are highlighted with a green box.

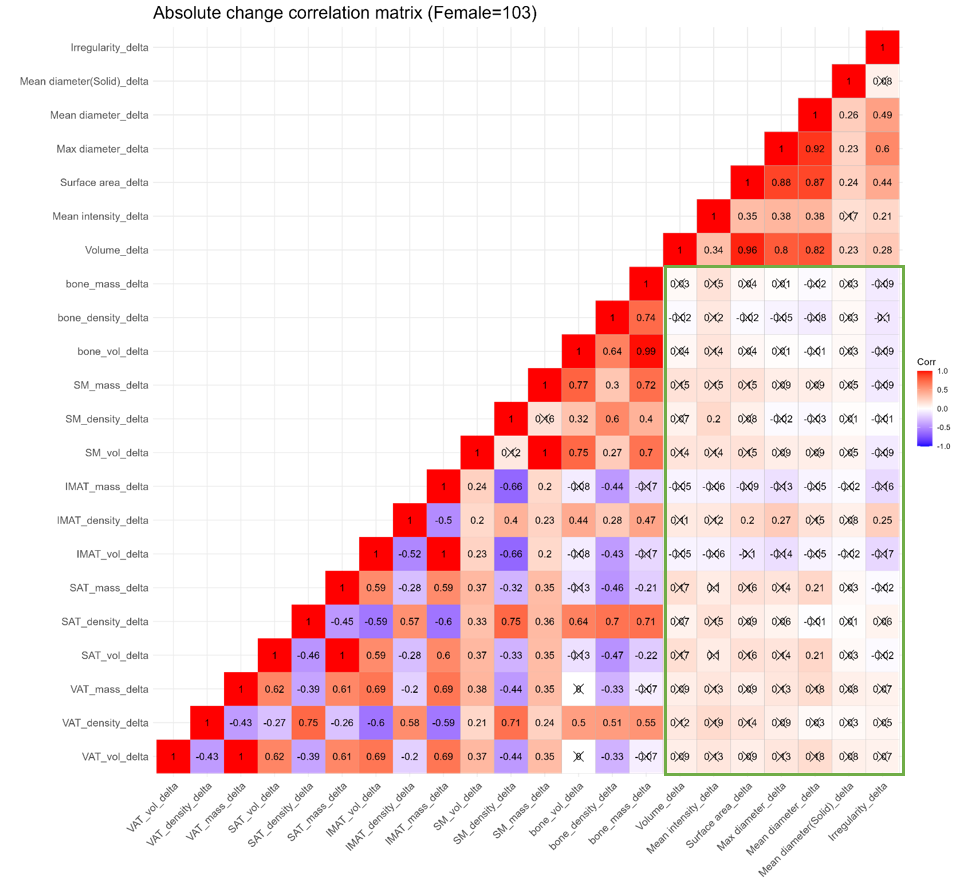

**S. Figure 5.** Correlation visualization between changes of body composition and nodule features (Female=103). Correlations marked with a cross are statistically insignificant (p-value > 0.05). The correlations between nodule and body composition features are highlighted with a green box.
